# Severe COVID-19 patients have impaired plasmacytoid dendritic cell-mediated control of SARS-CoV-2-infected cells

**DOI:** 10.1101/2021.09.01.21262969

**Authors:** Manon Venet, Margarida Sa Ribeiro, Elodie Décembre, Alicia Bellomo, Garima Joshi, Marine Villard, David Cluet, Magali Perret, Rémi Pescamona, Helena Paidassi, Thierry Walzer, Omran Allatif, Alexandre Belot, Sophie Assant, Emiliano Ricci, Marlène Dreux

## Abstract

Type I and III interferons (IFN-I/λ) are key antiviral mediators against SARS-CoV-2 infection. Here, we demonstrate that plasmacytoid dendritic cells (pDCs) are the predominant IFN-I/λ source following their sensing of SARS-CoV-2-infected cells. Mechanistically, this short-range sensing by pDCs requires sustained integrin-mediated cell adhesion with infected cells. In turn, pDCs restrict viral spread by an IFN-I/λ response directed toward SARS-CoV-2-infected cells. This specialized function enables pDCs to efficiently turn-off viral replication, likely *via* a local response at the contact site with infected cells. By exploring the pDC response in SARS-CoV-2 patients, we further demonstrate that pDC responsiveness inversely correlates with the severity of the disease. The pDC response is particularly impaired in severe COVID-19 patients. Overall, we propose that pDC activation is essential to control SARS-CoV-2-infection. Failure to unfold this response could be key to understand severe cases of COVID-19.

## Introduction

The innate immune system acts as the first line of defense for the sensing of viral infection. This involves rapid recognition of pathogen-associated molecular patterns (PAMPs), including viral nucleic acids, by pattern recognition receptors (PRRs). This recognition results in an antiviral response characterized by the production of type I and III (λ) interferons (IFN) and other pro-inflammatory cytokines, along with the expression of IFN-stimulated genes (ISGs). Whilst type I and III/λ IFNs interact with distinct receptors, they both induce similar signaling pathways and effector factors, thus referred herein to as the IFN-I/λ response^**1,2**^. This host response suppresses viral spread by blocking the viral life cycle at multiple levels, hereby promoting virus clearance. The IFN-I/λ response also mediates immunomodulatory effects in surrounding tissues and imparts the onset of the adaptive immune response^**3,4**^.

Severe acute respiratory syndrome coronavirus-2 (SARS-CoV-2) emerged in December 2019 and is responsible for the still-ongoing coronavirus disease 2019 pandemic^**5**^. Although most SARS-CoV-2-infected individuals experience asymptomatic to mild disease, others develop respiratory distress syndrome that is lethal in the most severe cases. Importantly, the IFN-I/λ response is now thought to be a critical host response against SARS-CoV-2 infection and its pathogenesis^**2,6-10**^. Recent studies have shown that SARS-CoV-2 can evade from the initial control by the IFN-I/λ response *via* manifold inhibitory mechanisms interfering with both the sensing and the signaling pathways within infected cells (review articles^**2,6,11,12**^ and illustrated in previous works^**10, 13-17**^). This immune evasion might lead to an increased viral load, followed by widespread inflammation^**6**^. Individual immune responses against viral infection can thus be extremely heterogenous, ranging from robust and fast IFN-I/λ production to impaired IFN-I/λ-mediated immunity. IFN-I/λ response is thus pivotal for the host defense against respiratory infections. Timing of the IFN-I/λ production also plays a crucial role in related Coronaviruses (*e*.*g*., MERS-CoV and SARS-CoV-1)^**18**^. Delayed IFN-I/λ signaling is associated with robust virus replication and promotes the accumulation of pathological monocyte-macrophages^**18**^. This results in lung immunopathology, vascular leakage and suboptimal T cell responses. Along this line, early reports on SARS-CoV-2 suggest that severe COVID-19 featured low level of IFN-I/λ but overproduction of inflammatory cytokines^**7,19-21**^. Accordingly, genetic deficiency, neutralization by autoantibodies directed against the IFN−I system, or viral-mediated inhibition of the IFN-I/λ response aggravates SARS-CoV-2 pathogenesis^**21-27**^. It is therefore critical to understand the regulation of the optimal production and activity of IFN-I/λ.

The plasmacytoid dendritic cells (pDCs) are a unique immune cell type specialized for rapid and massive production of IFN-I/λ^**28**^. pDCs possess multiple adaptations to efficiently produce IFN-I/λ^**28-32**^. As pDCs are refractory to most viral infections, their response is not directly repressed by viral proteins. This particularity contributes to the exceptional magnitude of pDC IFN-I/λ production^**28,33**^. The main viral-sensors responsible for pDC activation are Toll-like receptors (TLR)-7 and -9 that recognize viral RNA and DNA, respectively^**28**^. A recent report suggested that pDCs activated by SARS-CoV-2 differentiate into cytokine- and IFN-secreting effector cells, in a TLR-dependent manner^**34**^. Studies on related coronaviruses have demonstrated that pDCs migration into the lungs, and their rapid production of IFN-I is essential for the control of lethal infections by these coronaviruses^**19,35,36**^. Nonetheless, how pDCs respond to SARS-CoV-2-infected cells and how this response correlates with the progression of COVID-19 severity are still open questions.

Here, we explored the molecular mechanisms underlying the IFN-I/λ response against SARS-CoV-2 infection. Our results uncovered that pDCs establish cell contact with SARS-CoV-2 infected cells *via* α_L_β_2_ integrin/ICAM-1 adhesion complex and regulators of actin network. This physical contact between pDCs and infected cells is required for the pDC-mediated antiviral response by TLR7 recognition. Capitalizing on our findings that pDCs strongly respond by physical sensing of SARS-CoV-2-infected cells, we then showed that impaired pDC IFN-I/λ response associates with COVID-19 severity. It is now increasingly recognized that pDCs differentiate into different subsets with distinct phenotypes and functionalities. Here, we showed that the differentiation of pDCs into subsets is altered when stimulated by contact with SARS-CoV-2-infected cells as compared to other activation. Especially, when in contact with SARS-CoV-2-infected cells, pDCs preferentially differentiate into a subset, which efficiently produces IFN-I/λ, then leading to a robust antiviral control directed towards the infected cells.

## Results

### Robust activation of pDCs in response to SARS-CoV-2-infected cells

Respiratory epithelial cells represent the first infected tissue in the course of SARS-CoV-2 infection. To investigate which hematopoietic cell type is primarily responsible for the IFN-I/λ response against SARS-CoV-2 infection, human peripheral blood mononuclear cells (PBMCs) isolated from healthy donors were cocultured with SARS-CoV-2-infected human lung-derived cells. Calu-3 cells and A549-ACE2 cells (expressing the angiotensin-converting enzyme 2) were infected for 48 hours prior to coculture with PBMCs (including pDCs) for 16-18 hours. We found that PBMCs respond by a robust secretion of IFNα when cocultured with SARS-CoV-2-infected cells (**Fig. 1a-b**). In contrast, the supernatants (SN) of SARS-CoV-2-infected cells failed to trigger IFNα secretion by PBMCs (**Fig. 1a-b**). As expected from previous publications, SARS-CoV-2-infected cells did not produce themselves detectable level of IFNα^**19,37-42**^. Plasmacytoid dendritic cells (pDCs) are known to robustly produce IFN-I/λ, notably IFNα ^**28**^. In line with this, antibody mediated-depletion of pDCs from PBMCs abolished IFNα secretion in response to co-cultured SARS-CoV-2-infected cells (**Fig. 1a-b**). To further demonstrate that pDCs are the major source of IFNα upon incubation with SARS-CoV-2-infected cells, pDCs were purified from PBMCs. Purified pDCs potently produced IFNα in response to co-culture with SARS-CoV-2-infected cells (**Fig. 1a-b)**.

**Fig. 1.**
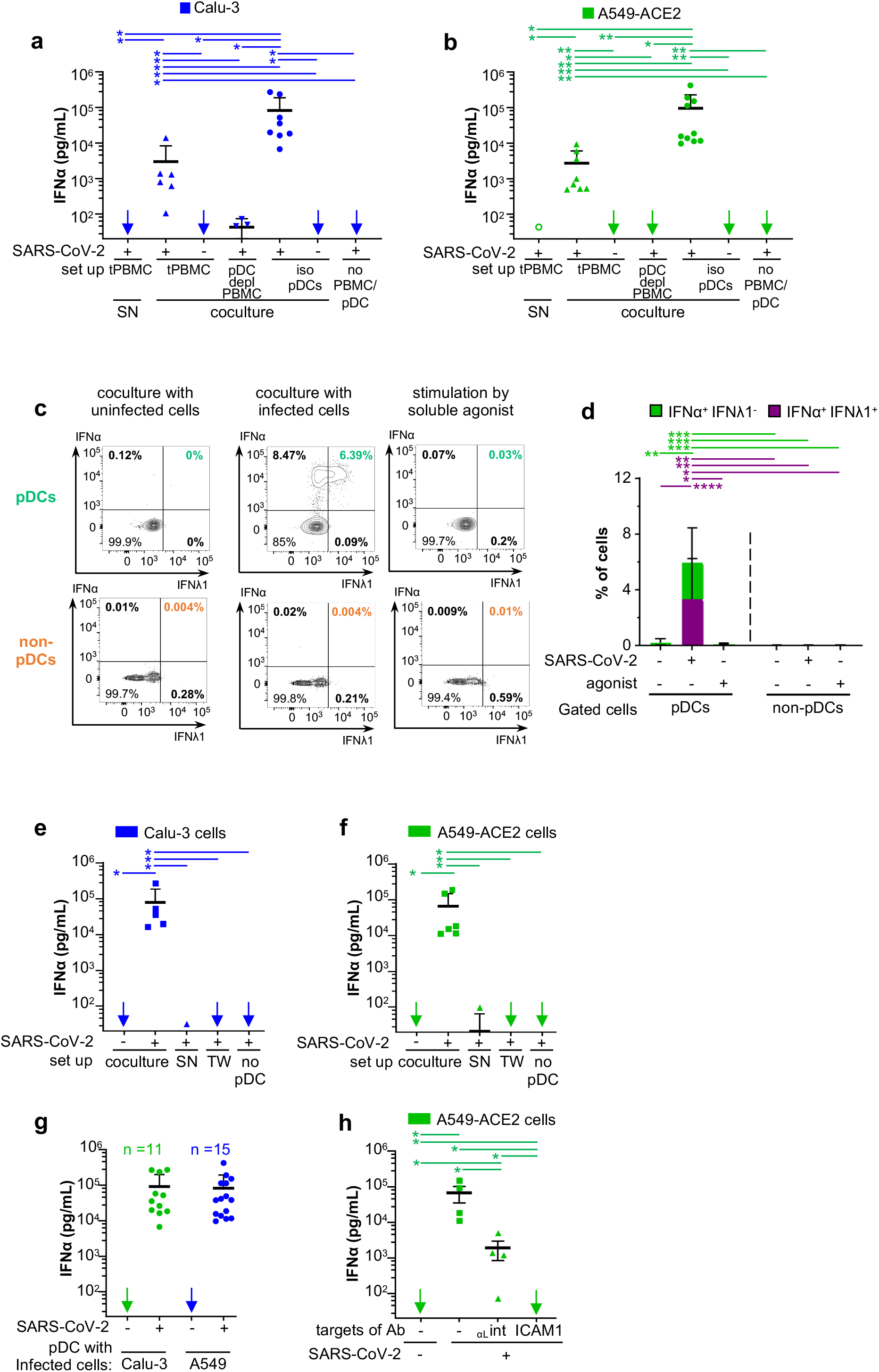
pDCs are the main IFN*α* producers in response to contact with SARS-CoV-2-infected cells. Cells were infected by SARS-CoV-2 (indicated as +) or not (indicated as -) two days prior to coculture with PBMCs, pDC-depleted PBMCs and isolated pDCs. Infected cell types included the human alveolar basal epithelial cell lines *i*.*e*., Calu-3 and A549-ACE2. Statistical analyses of the results were performed using Wilcoxon rank-sum test and and p-values were calculated with Tukey and Kramer test. The significant contrasts are indicated when p-values are: ≤0.05; * and ≤0.005; **. **a-b**, Quantification of IFNα in the supernatants of PBMCs (tPBMC), pDC-depleted PBMCs (pDC depl PBMC) and isolated pDCs (iso pDC) cocultured with either SARS-CoV-2-infected or uninfected Calu-3 cells (**a**) or A549-ACE2 cells (**b**), or treated with 100*µ*l of cell-free supernatant (SN) collected from SARS-CoV-2-infected cells. Of note, viral titers of SARS-CoV-2: SN≈2.5×10^5^ foci forming units (ffu)/ml, MOI ≈ 1/pDCs. Arrows indicate results below the detection threshold of the IFNα ELISA (*i*.*e*., 12.5 pg/ml). Each dot represents one independent experiment (n=4-9) performed with distinct healthy donors (ELISA results are similarly presented in all other Figures). Error bars represent the means ± standard deviation (SD); ind./cocult. = induction/coculture. **c-d**, Total PBMCs were cocultured with SARS-CoV-2-infected or uninfected A549-ACE2 cells or treated with TLR agonists [31.8 *µ*M R848 and 42.22 *µ*M polyI:C] for 14 to 16 hours. Cell populations gated as pDC and non-pDC PBMCs (*data not shown*). Representative dot blot of flow cytometry analyses (**c**) and frequencies of cells positive for IFNα^+^ and/or IFNλ1^+^ in gated pDCs (left bars) *versus* non-pDC PBMCs (right bars) (**d**). Means ± SD; Dots represent n=10-11 independent experiments/distinct healthy donors. **e-h**, Quantification of IFNα in SNs of pDCs cocultured with the indicated cell types infected or not by SARS-CoV-2. **e-f**, pDCs were cocultured with infected cells, either in direct contact (coculture) or physically separated by the semi-permeable membrane of transwell (TW), or treated with SN from the corresponding SARS-CoV-2-infected cells. IFNα concentration was also determined in the SN of SARS-CoV-2-infected cells cultured without pDC (no pDC). Means ± SD; n=3-9 independent experiments. **g**, Dots represent for pDCs purified from distinct donors in independent experiments; including n=11 and n=15 for A549-ACE2 and Calu-3 cells, respectively. Means ± SD; **h**, Quantification of IFNα in SN of pDCs cocultured with SARS-CoV-2-infected cells A549-ACE2 treated or not with blocking antibodies against α_L_-integrin and ICAM-1 at 10 *µ*g/mL. Means ± SD; dots represent n=4-5 independent experiments.

Consistent with these results, we further showed that IFNα producer cells were markedly enriched in the cell population gated as pDCs as compared to other hematopoietic cell types (*i*.*e*., non-pDCs): no IFNα^+^ cells detected among non-pDCs **(Fig. 1c-d)**. Most IFNα^+^ pDCs concomitantly produced IFNλ in response to SARS-CoV-2-infected cells (**Fig. 1c-d)**. In contrast, stimulation by soluble agonist elicited IFNλ^+^ pDCs, but no detectable IFNα^+^ cells (**Fig. 1c-d)** and a potent upregulation of surface expression of activation markers including CD83 and the programmed cell death ligand-1 (PD-L1) as compared to coculture with SARS-CoV-2-infected cells (*data not shown*). This suggested that the pDC response to SARS-CoV-2-infected cells is likely qualitatively distinct from stimulation by soluble agonist. CD83 and PD-L1 are induced by NF-κB-signaling, and thus dependent on signaling distinct from the IFN-I/λ response^**43,44**^.

Next, we tested whether the sensing of SARS-CoV-2-infected cells required a productive infection of pDCs, using a recombinant SARS-CoV-2 infectious clone expressing the mNeongreen reporter (icSARS-CoV-2-mNG^**45**^). No infection of pDCs (defined as CTV^+^-pDCs) was detected when pDCs were incubated in contact with icSARS-CoV-2-mNG-infected A549-ACE2 cells (*data not shown*). However, in the same coculture set up, infection by icSARS-CoV-2-mNG was readily detected in initially uninfected/naive RFP^+^ A549-ACE2 cells, hence proving validation of efficient viral transmission.

Taken together, these results strongly suggest that IFNα is robustly produced only by pDCs. This occurs by sensing of SARS-CoV-2-infected cells without productive infection and likely induces a specific activation state in pDCs.

### Short-range sensing of infected cells *via* cell-contact triggers the pDC IFNα response

We observed that cell-free SN from SARS-CoV-2-infected cell types failed to trigger IFNα production by PBMCs or by purified pDCs (**Fig. 1a-b)**. To avoid possible misinterpretation due to contamination by cell debris and/or floating cells^**34**^, we selected an incubation time for SARS-CoV-2 infection so that no cytolytic effect was detectable in infected human lung-derived cells when cells/SNs were collected for coculture. Nevertheless, no pDC activation by cell-free SN was readily detected even at a multiplicity of infection (MOI) of 5 per pDC. To further determine if cell-to-cell contacts were required for the transmission of the immunostimulatory signal to pDCs, we used transwell chambers containing SARS-CoV-2-infected cells and pDCs separated by a 0.4μm permeable membrane. This physical cell separation fully prevented IFNα production by pDCs (**Fig. 1e-f**). To confirm that this feature was not cell type specific, we used a variety of lung-derived cell lines Calu-3, A549-ACE2, H358-ACE2 as well as non-lung Huh7.5.1 cells. We found that pDCs were not stimulated by cell-free virus produced by any of these cells not in presence of infected cells but physically separated, while they were activated by all tested SARS-CoV-2-infected cells (**Fig. 1e-f**). Remarkably, similar levels of IFNα secretion were reproducibly obtained for pDCs isolated from several distinct healthy donors (**Fig. 1g**; as n=11 and n=15 healthy donors for A549-ACE2 and Calu-3 cells, respectively).

To further define the mechanisms underlying pDC activation upon contact with SARS-CoV-2-infected cells, we assessed the implication of cell adhesion complexes in this process. We focused on α_L_ integrin and its ligand Intercellular Adhesion Molecule (ICAM)-1 (also called CD54) ^**46**^, respectively, highly expressed by pDCs and by various cell types, guided by previous studies on the regulation of pDCs in the context of other infections ^**47**^. Antibody-mediated blockade of both α_L_ integrin and ICAM-1 greatly prevented pDC IFNα production (**Fig. 1h** and *data not shown*). The engagement of integrins by their ligands is known to induce local recruitment of the actin network. Notably Arp2/3 complex mediates actin nucleation by recruiting and branching actin filaments within the network^**48-51**^. Pharmacological inhibition of Arp2/3 complex impaired IFNα production by pDCs in coculture with SARS-CoV-2-infected cells, in a dose dependent-manner (*data not shown*). This suggested that cell adhesion-induced actin recruitment is likely involved in the structuration of cell contacts. Furthermore, using specific TLR7 antagonist (*i*.*e*., IRS661), we showed that the endosome-localized TLR7 sensor mediates the sensing of SARS-CoV-2-infected cells by pDCs (*data not shown*).

Together our results demonstrated that physical contact between pDCs and SARS-CoV-2-infected cells is required for pDC IFNα production. This contact involves cell adhesion mediated by α_L_ integrin/ICAM-1 complexes, which likely remobilize the actin network at the cell-to-cell contact, and leads to a robust TLR7-dependent IFNα response by the pDCs.

### IFN-I/λ signature in patients at early time-point of SARS-CoV-2 infection

Based on the findings that pDCs are the main cell type producing IFNα in response to SARS-CoV-2-infected cells, we sought to explore how this singular activation mechanism for IFN-I/λ response is modulated in the course of the infection in patients and how it could relate to COVID-19 severity. A longitudinal study of IFN-I/λ response was done with different subsets of patients: *i)* critically ill, herein referred to as *Severe* patients, who presented acute respiratory distress syndrome or severe pneumonia at hospital admission and required mechanical ventilation in intensive care units, and *ii)* patients with mild symptoms (*i*.*e*., low-grade fever, cough, malaise, rhinorrhea, sore throat), that group was sub-divided according to the days of sample collection post-symptom onset, *i*.*e*., *Mild early* for the first two weeks and *Mild late* at the later time points. Of note, most patients were SARS-CoV-2 positive in nasal swab samples by qPCR at sampling time in the *Mild early* group but not anymore or presenting low viral levels in the *Mild late* group. All patients and the analyzed time-points are listed (*data not shown*), which provides information on the clinic and viral loads in nasal swab samples.

We quantified the IFN-I/λ levels in blood samples of infected patients at both transcriptional and protein levels for secreted IFNα, IFNλ1, IFNγ and IL-6. Both approaches revealed an elevated IFN-I/λ response at early time points (within the first 10-11 days post-onset of symptoms) for all patients with mild symptoms, that seemed to vanish over time, mirroring the controlled decrease of the viral load (**Fig. 2a**). The IFN-I/λ response was elevated in *Severe* patients even at late time-points. Of note, the pro-inflammatory cytokine IL6 was greatly detected in *Severe* patients as compared to other groups (**Fig. 2a**). This positive correlation of the inflammatory response with COVID-19 severity is in agreement with previous reports^**7,52-54**^.

**Fig. 2.**
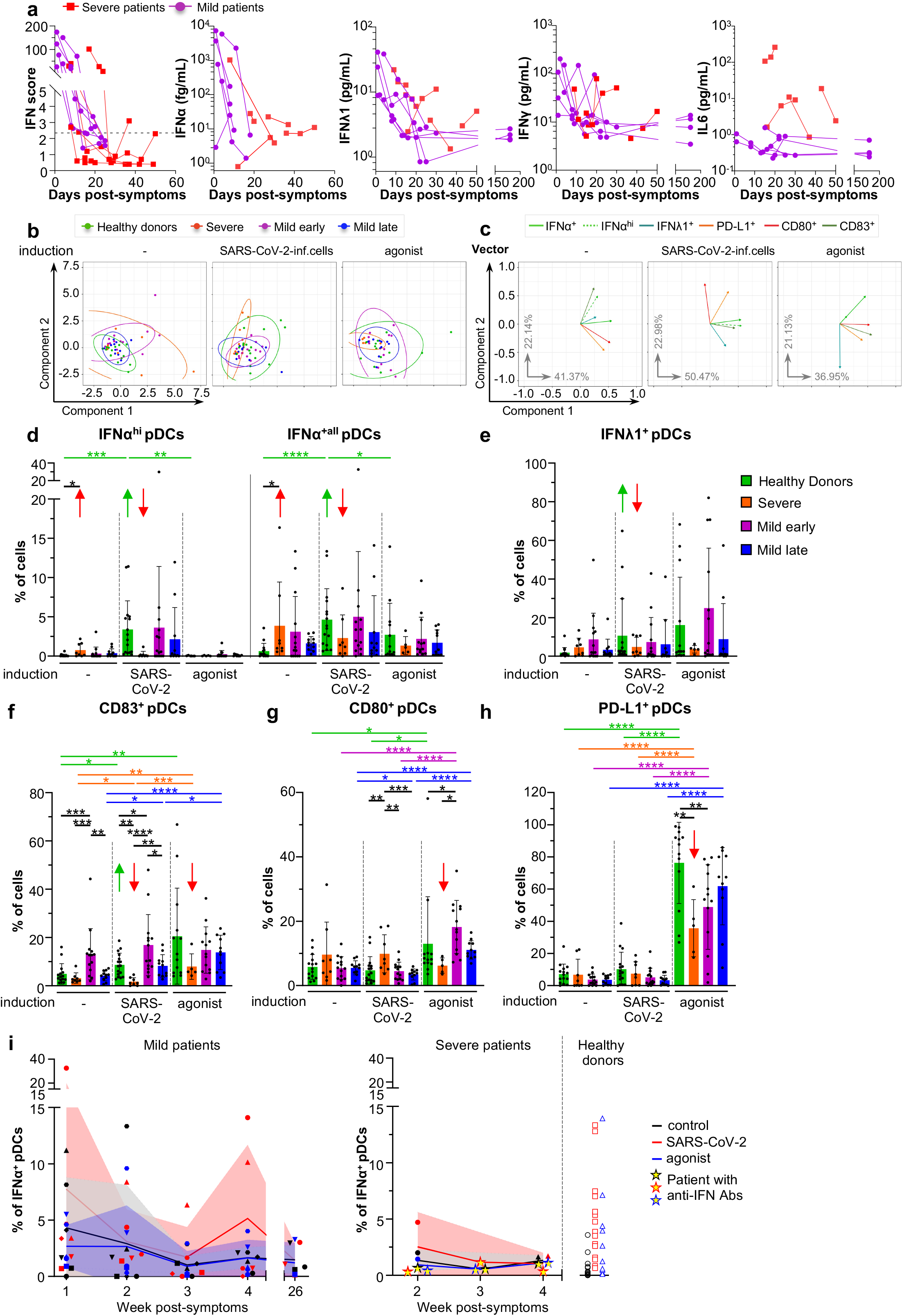
Innate responses against SARS-CoV-2-infected analyzed for patients with various COVID severities. **a**, Kinetic analysis of cytokine profile of *Severe* and *Mild* COVID-19 patients. Determination of the IFN score in whole blood as well as secreted IFNα, IFNλ1 and IFNγ in plasma, at the indicated time-points. **b-i**, PBMCs issued from the indicated groups of patients (*i*.*e*., *healthy donors, Severe, Mild early, Mild late*, see description of the patients in Material and Methods section) were cocultured for 14-16 hours with SARS-CoV-2-infected or uninfected A549-ACE2 cells, or treated with agonists [31.8 *µ*M R848 and 42.22 *µ*M polyI:C or 2.04 *µ*M LPS]. **b-c**, PCA analyses of the quantification by multiparametric flow cytometry in cell populations gated as pDCs (*data not shown*). Clustering of the groups of patients was performed by combining the detection of multiple cell surface expressed-differentiation markers and intracellular cytokines. Each dots represent one individual patient sample (**b**) and the eigen vectors that represent the first and second main components, with indication of the percentage of the variance explained by each principal component (**c**). **d-h**, Quantification of the frequency of cells positive for IFNα^hi^/IFNα^+all^ (**d**) IFNλ1^+^ (**e**) and the differentiation markers, CD83, CD80, PD-L1 (**f-h**) in gated pDCs. **i**, Kinetic analysis of the ability of pDCs from *Mild* (left panel), *Severe* patients (middle panel) and *healthy donors* (right panel) to mount an IFNα^+^ response upon *ex vivo* stimulation with SARS-CoV-2-infected cells (red) agonist (blue) *versus* control cells (black). Dots for the *Severe* patient with circulating anti-IFNAR antibodies (*data not shown*) are represented by yellow-centered stars. Patient PBMCs were collected from the symptom onset and results correspond the timeframes as follows defined in weeks: 1= [Days 1-8]; 2= [Days 8-15]; 3= [Days 15-22]; 4= [Days 22-30]. Means (colored lines) and errors (colored areas) are indicated (n=20-24 analysed patients).

We then determined the ability of PBMCs isolated from the different groups of patients to respond to *ex vivo* stimulation by SARS-CoV-2-infected cells. As our results pointed out the importance of pDCs in the IFN-I/λ response mounted against SARS-CoV-2-infected cells, we primarily analyzed pDCs. This response was compared to the one induced by TLR7 [R848]/TLR3[polyI:C] agonist. A group of *healthy* donors comprising similarly treated samples was used as reference. Post-stimulation, we performed a multiparametric flow cytometry analysis to define the profiles of cell surface expression of activation markers and intracellular cytokines. These expression profiles were further assigned to different pivotal innate immune cells by designing a panel of cell type markers (*data not shown*).

Collectively, our results demonstrated distinct cytokine profiles across COVID-19 severities, and allow to assign patient groups and biomarkers suitable for a functional analysis of the cell responsiveness in *ex vivo* stimulation.

### COVID-19 severity correlates with a blunted pDC IFNα response to SARS-CoV-2-infected cells

Principal component analysis (PCA) of the flow cytometry datasets was performed within gated pDCs and for each stimulation condition to test whether differences in activation and cytokine expression could be linked to COVID-19 severity (**Fig. 2b-c**). PCA allows to reduce high dimensional data into principal components that maximize the variance of the projected data and potentially identify the underlying factors associated with it. Each patient is placed with respect to its distribution in the first two principal components (PCs), which explain most of the data variance (**Fig. 2b**), and vectors corresponding to the eigen decomposition of the data covariance matrix are depicted to identify features that explain the observed variance (**Fig. 2c**). For pDCs analyzed in the unstimulated condition, the first two principal components (PCs) revealed a differential segregation of the patients: the *Severe* group (in red), and to some extent *mild early* group (in purple) were mainly spread across the first PC (which was explained by IFNα, CD80 and PD-L1), while *healthy* and *Mild late* individuals were tightly clustered in both PCs (**Fig. 2b-c**, left panels). An opposite segregation of the patient groups was observed in pDCs when PBMCs were cocultured with SARS-CoV-2-infected cells (**Fig. 2b-c**, middle panels). Especially, this stimulation induced a spread across the first PC (mainly driven by IFNα/IFNα^hi^ and CD83 expression) of patients from the *Mild* and *healthy* groups, in accordance with the pDC ability to respond to SARS-CoV-2-infected cells (**Fig. 2c**, middle panel). Of note, the *healthy* individuals were spread across the two PCs, whereas the *Mild early and Mild late* groups dispersed primarily along the first PCs. On the contrary, individuals from the *Severe* group were spread across the second PC (driven by CD80 and PD-L1 expression). Regarding pDC response to synthetic agonist, all groups were similarly spread across the first and second PCs, while the *Severe* group was tightly grouped within the second PC (driven by IFNλ expression) (**Fig. 2b-c**, right panels). These results suggested that pDCs from *Severe* individuals display differences in activation markers and cytokine expression (*i*.*e*., IFNα, CD80, CD83) compared to individuals from all the other groups. These differences were particularly observed in absence of stimulation and upon stimulated by SARS-CoV-2-infected cells.

We further analyzed the expression of activation markers and cytokines individually at the single-cell level, focusing first on markers identified as driving forces in the PCA analyses. Similar to **Fig. 1**, the frequency of IFNα producer pDCs greatly increased in response to SARS-CoV-2-infected cells in the *healthy* donor group (**Fig. 2d**, green arrows). In sharp contrast, pDCs from *Severe* patients failed to be activated by SARS-CoV-2-infected cells as revealed by the absence or low detection of IFNα, IFNλ and CD83 (**Fig. 2d-f**; red bars and arrows). Of note, a strong basal level of pDC IFNα expression was observed in the absence of *ex vivo* stimulation for the *Severe* patients (using less stringent discriminative gating of positive pDCs, noted distinctively as to IFNα^+all^; **Fig. 2d**). The response to agonist stimulation was also greatly diminished in pDCs from *Severe* patients compared to *healthy* donors and patients with *mild* symptoms, notably as shown by the level of activation markers (CD83, CD80 and PD-L1; **Fig. 2f-h**; red arrows). Overall the results obtained by analyses of the markers individually were in agreement with the PCA data, since the major change in pDC responsiveness concerned *Severe* patients as compared to the other groups. As highlighted in the identified first PCs of PCA analysis, this difference was primarily explained by lack of IFNα and CD83 expression in pDCs upon stimulation by SARS-COV-2-infected cells.

In addition to pDCs, other cell types were also selected for multiparametric flow cytometry analysis based on their potential to play pivotal first defense against viruses. Especially, the Th1-promoting myeloid/conventional dendritic cell subset (mDC1), which produces IFNλ *via* TLR3-mediated recognition of viral RNA and the Th2-promoting myeloid/conventional DC subset (mDC2) and monocytes *i*.*e*., Human Leukocyte Antigen – DR isotype (HLA-DR)^+^CD14^+^ populations, known to produce proinflammatory cytokines^**55,56**^. In line with the results obtained with pDCs, expression of IFNλ1 was barely detectable in the mDC1 subset of *Severe* patients upon stimulation by SARS-CoV-2-infected cells and agonists, comparatively to the other groups (*data not shown*). As expected, in other cell population *(i*.*e*., mDC1, mDC2, non-mDC2 and HLA-DR^+^CD14^+^ populations), other markers (*i*.*e*., IFNα, IL6, CD83, CD80 and PD-L1) were not readily induced by SARS-CoV-2-infected cells even in *healthy* donors (data not shown). Nevertheless, our data showed a potent upregulation of IL6 by HLA-DR^+^CD14^+^ subset upon agonist treatment in *Severe* patients and *Mild early* (*data not shown*). HLA-DR^+^CD14^+^ population represents a highly frequent cell subset of non-mDC2 (*e*.*g*., among the gated live cells^+^ lin^-^ HLA-DR^+^: 65.7% and with exclusion of the few mDC2). In accordance, a high frequency of IL6^+^ cells was also observed for the non-mDC2 populations (*data not shown*). Likewise, *Severe* patients presented an elevated level of blood IL6 (**Fig. 2a**). These results are consistent with the previously reported production of pro-inflammatory cytokines by monocytic cells in patients with severe disease^**7,53**^, and provided further insights into the differential responses of other immune cells.

### Dynamics of the IFN-I/*λ* response in COVID-19 patients

Next, we sought to define the dynamics of the response in COVID-19 patients. First, kinetic analyses in *Mild* patients revealed an elevated frequency of pDC IFNα^+^ levels in absence of stimulation that vanished over time while, relatively, their ability to respond to SARS-COV-2-infected cells increased at late time-points (**Fig. 2i**, left panel). A limited induction of pDC IFNα^+^ in response to SARS-COV-2-infected cells was observed in *Severe* patients at all analysed time-points (**Fig. 2i**, right panel). We next performed a kinetic analysis for a *Severe* patient, who had high level of anti-IFNα antibody detected in the blood (*data not shown*)^**21-27**^. This analysis showed that the ability of pDC to respond to stimulation was blunted in this patient (**Fig. 2i**, right panel; represented by stars with yellow-center). The kinetic of pDC IFNλ^+^ response presented a similar pattern for both *Mild* and *Severe* groups with basal level in absence of stimulation at early time-points and low response to SARS-CoV-2 infected cells, while pDC response recovered at late time, and this was paralleled by the IL6^+^ frequency in HLA-DR^+^CD14^+^ population (*data not shown*).

Together our results demonstrated that the monocytic subsets likely contribute to an exacerbated pro-inflammatory response implying notably IL6 production, but that is liklely not triggered directly by the contact with SARS-CoV-2-infected cells. As opposed, impaired IFN-I/λ response following cell contact between SARS-CoV-2-infected cells and pDCs from *Severe* patients, including those with anti-IFNα antibodies, suggested a ‘*silencing/unresponsive state*’ of pDCs in this context. This might be due to the lack of an amplification-loop by ISG resulting in lower activated state and IFNα production by pDCs and to some extent, similarly for IFNλ1 expression by mDC1.

### Limited pDC differentiation and cytotoxic activity when in contact with SARS-CoV-2-infected cells

As we found that pDC activation is a salient feature that negatively correlates with COVID-19 severity, we aimed to determine how the contact with SARS-CoV-2-infected cells impacts the varied downstream signaling and function of pDCs. First, we analyzed the expression of pDC surface molecules enabling the stimulation of adaptive responses, namely HLA-DR, an MHC class II cell surface receptor driving the activation of CD4^+^ T cells, and the B cell ligand CD70 expressed by pDCs and known to interact with CD27 and to induce proliferation and differentiation of B cells into plasmablast^**57**^. Our results showed that pDCs upregulated both surface molecules in response to contact with SARS-CoV-2-infected cells, although at a lower level compared to stimulation by cell-free virus (influenza virus; flu) and synthetic agonists (**Fig. 3a-b**).

**Fig. 3.**
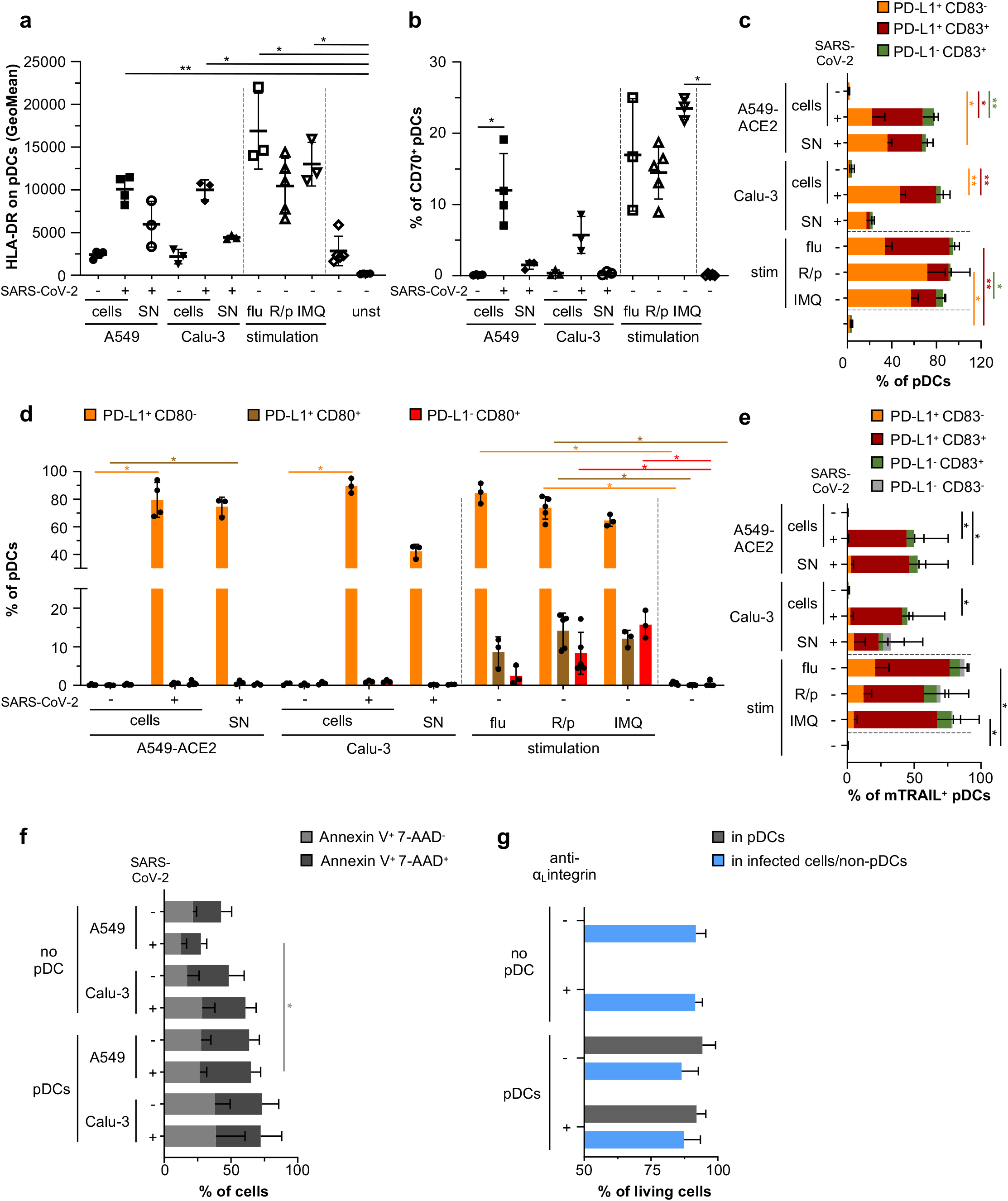
SARS-CoV-2-infected cells induce pDC maturation and phenotypic diversification. Human pDCs isolated from healthy donors were cocultured with SARS-CoV-2-infected (+) or uninfected (-) A549-ACE2 or Calu-3 cells (indicated as ‘cells’), or were stimulated with 100*µ*l of cell-free SN collected immediately prior coculture from the corresponding SARS-CoV-2-infected cells (viral titers ≈ 2.5×10^5^ ffu/ml -MOI ≈1/pDCs), or were stimulated with influenza virus (flu; viral titers ≈ 10^7^ pfu/ml -MOI ≈ 0.5/pDCs), R848/polyI:C (R/p) or imiquimod (IMQ) for 14-16 hours (stim = stimulation; unst = unstained). **a-e**, Quantification by flow cytometry of HLA-DR Geomean (**a**) or the frequency of pDCs positive for CD70 (**b**), PD-L1 and/or CD83 (**c**), PD-L1 and/or CD80 (**d**), PD-L1 and/or CD83 among mTRAIL^+^ pDCs (**e**) determined in gated pDCs (live cells^+^ singulets^+^ CD123^+^ BDCA2^+^). **f**, SARS-CoV-2-infected (+) or uninfected (-) A549-ACE2 or Calu-3 cells were cultured alone (no pDC) or cocultured with pDCs for 14-16 hours. Quantification by flow cytometry of the frequency of gated A549-ACE2 and Calu-3 cells positive for Annexin V and/or 7-AAD. **g**, icSARS-CoV-2-mNG-infected A549-ACE2 were cultured alone (no pDC) or cocultured with pDCs in the presence or absence of anti-_αL_integrin blocking antibody for 48 hours. Quantification by flow cytometry of the frequency of living cells using live-dead marker in the gated pDCs (stained with CellTraceTM Violet prior to coculture) and infected cells/non-pDC. Bars represent means ± SD; n=3-5 independent experiments using distinct healthy donors. Each dot in panels **a-b, e**, represents one independent experiment. The data were analyzed using Kruskal-Wallis Global test and p-values were calculated with Tukey and Kramer test; * ≤0.05 and ** ≤0.005.

Similarly, surface expression of CD83, an activation marker for antigen presenting cells^**58,59**^ was induced jointly with PD-L1 upon contact with SARS-CoV-2-infected cell (**Fig. 3c**). SN from SARS-CoV-2-infected cells induced HLA-DR, CD83 and PD-L1 expression, but not as potently as other soluble agonists. These results obtained with purified pDCs are in agreement with results obtained with pDCs present in PBMCs cocultured with SARS-CoV-2-infected cells (**Fig. 1)**.

pDCs are now recognized to be a heterogeneous population composed of subsets endowed with diversified functions^**28,34,60-63**^. Importantly, stimulation of pDCs can impact the frequency and phenotype of these diversified subsets. We thus assessed the expression of a set of surface molecules previously assigned to define specific subpopulations of pDCs, *i*.*e*., CD2, CD5, AXL, CD80 and PD-L1^**28,34,60-63**^. CD2^hi^ pDCs have a survival advantage and are able to efficiently trigger proliferation of naive allogenic T cells^**61,62,64**^. Stimulation of pDCs by contact with SARS-CoV-2-infected cells did not impact the frequency of CD2^hi^ pDCs, and this CD2^hi^ subset displayed an activation profile similar to the one of CD2^low^ subset (*data not shown*). CD2^hi^CD5^+^AXL^+^ pDCs were defined as a subset that display limited IFN-I production capacity but can potently activate T cells, but represent a very scarce subpopulation of pDCs^**63**^. The CD2^hi^CD5^+^AXL^+^ pDCs modestly decreased upon stimulation by SARS-CoV-2-infected cells, their SN or TLR-agonists, yet whether this intriguing decrease is relative to the activated state of pDCs requires further investigation (*data not shown*). We then addressed the diversification of pDCs into functionally distinct populations defined by PD-L1/CD80 expression. In agreement with previous reports, the stimulation by cell-free agonists and influenza virus triggered the differentiation into all subsets: PD-L1^+^CD80^-^, PD-L1^+^CD80^+^ and PD-L1^-^CD80^+^ pDCs^**34,60**^. In sharp contrast, direct contact with SARS-CoV-2-infected cells restricted the differentiation of pDCs only towards the PD-L1^+^CD80^-^ subset (**Fig. 3d**).

Collectively, these results revealed that, unlike activation by cell-free viruses and agonists, direct activation of pDCs by SARS-CoV-2-infected cells restricted their differentiation into specific functional subsets, *i*.*e*., inducing a maturation only into PD-L1^+^CD80^-^ subset.

We observed that pDCs expressed PD-L1 (*i*.*e*., programmed cell death ligand-1) when cocultured with SARS-CoV-2-infected cells. Therefore, we further examined the induction of cytotoxic activity of pDCs. In keeping with the previously-reported induction of membrane-bound TNF-related apoptosis-inducing ligand (mTRAIL) expression by pDCs upon viral stimulation (*e*.*g*., HIV)^**65-67**^, we found that mTRAIL was readily co-expressed along with PD-L1 and CD83 upon stimulation by influenza virus and synthetic agonists (**Fig. 3e**). Nevertheless, mTRAIL upregulation was more limited upon pDC coculture with SARS-CoV-2-infected cells as compared to stimulation by cell-free stimulation (**Fig. 3e**, total % of the bars). Moreover, the frequencies of Annexin V^+^/7-ADD^+^ apoptotic Calu-3 and A549-ACE2 cells in cocultures with pDCs were similar between infected (*i*.*e*., pDC activation) and uninfected (*i*.*e*., no pDC activation) conditions, hence suggesting that mTRAIL expression on activated pDCs did not endow these cells with cytotoxic activity (**Fig. 3f**). In line with this, the contact with activated pDCs did not markedly impact the viability of the cocultured SARS-CoV-2-infected cells (*i*.*e*., when comparing condition with or without the anti-α_L_ integrin, which inhibits contact and pDC activation), nor the viability of activated pDCs themselves, even when analyzed after 48 hours of coculture (**Fig. 3g**).

Overall, these results showed that activation of pDCs by SARS-CoV-2-infected cells did not induce cytotoxic activity and led to their preferential diversification into functional pDC subsets known to be specifically able to robustly produce IFN-I/λ.

### pDC activation by SARS-CoV-2-infected cells primarily leads to the antiviral state *via* IFN-I/λ production

We then sought to examine deeper the signaling pathways at play in pDCs upon activation by coculture with SARS-CoV-2-infected cells. TLR7-dependent activation of pDCs can induce a ‘bifurcated’ signaling leading to *(1)* IFN-I/λ production mostly *via* IRF7-related signaling and other cytokines and *(2)* activation/differentiation markers and inflammatory cytokines *via* NF-κB-pathway^**28,68**^. We noticed that expression of the activation markers HLA-DR, CD70, CD83, mTRAIL by pDCs was weaker when induced by contact with SARS-CoV-2-infected cells compared to soluble agonist stimulation (**Fig. 3**). These markers/proteins are primarily regulated by NF-κB-mediated signaling (*data not shown*)^**43,44**^. To further define the signaling active in pDCs, we quantified transcripts levels of representative key effectors regulated by IRF/IFN-I signaling (*i*.*e*., *mxA, isg15* and *ifnλ) versus* those primarily regulated by NF-κB-pathway (*i*.*e*., *il-6* and *tnfα)* (*data not shown*). Cocultures of pDCs and SARS-CoV-2-infected cells induced more the IRF7/IFN-I-regulated molecules than the representatives of the NF-κB-pathway (**Fig. 4a**). This was again confirmed by the quantification of secreted cytokines in the cocultures, demonstrating higher levels of IRF7/IFN-I-regulated IFNλ1/2/3 compared to TNFα (**Fig. 4b** compared to **4c**). This observation was also in agreement with the high level of secreted IFNα in similar experiments (**Fig. 1a-b**). Of note, no detectable cytokine and low level of the corresponding transcript expression were detected in SARS-CoV-2-infected cells in the absence of pDCs, or when pDCs were stimulated by SARS-CoV-2 SNs. In these assays, the detection included expression by both pDCs and coculture infected cells. Thus, the low activation level of the NF-κB-pathway and/or its variability among different infected cell types can be explained by a contribution of the infected cells, as a feedback loop of the response to activated pDCs. Hence, we assessed cytokine expression at single-cell level by flow cytometry in the gated pDCs (**Fig. 4d**). Stimulation by SARS-CoV-2-infected cells elicited higher frequency of IFN*α*^+^ pDCs compared to TNF*α*^+^ pDCs (**Fig. 4d**) and again, this result contrasted with the limited frequency of IFN*α*^+^ pDCs in response to cell-free agonist. Remarkably, almost all TNF*α*^+^ pDCs were also IFN*α*^+^ when stimulated by contact with SARS-CoV-2-infected cells, as opposed to the detection of TNF*α*^+^IFN*α*^-^ pDCs not IFN*α*^+^ when stimulated by cell-free influenza virus **(Fig. 4d)**. This possibly implied that the activation of the NF-κB-pathway was subsequent to IRF7/IFN-I response when induced by SARS-CoV-2-infected cells. Similar analysis performed for IFN*α*^+^ combined with IFNλ1^+^ also revealed that virtually all IFNλ1^+^ pDCs were IFN*α*^+^ (**Fig. 4e**).

**Fig. 4.**
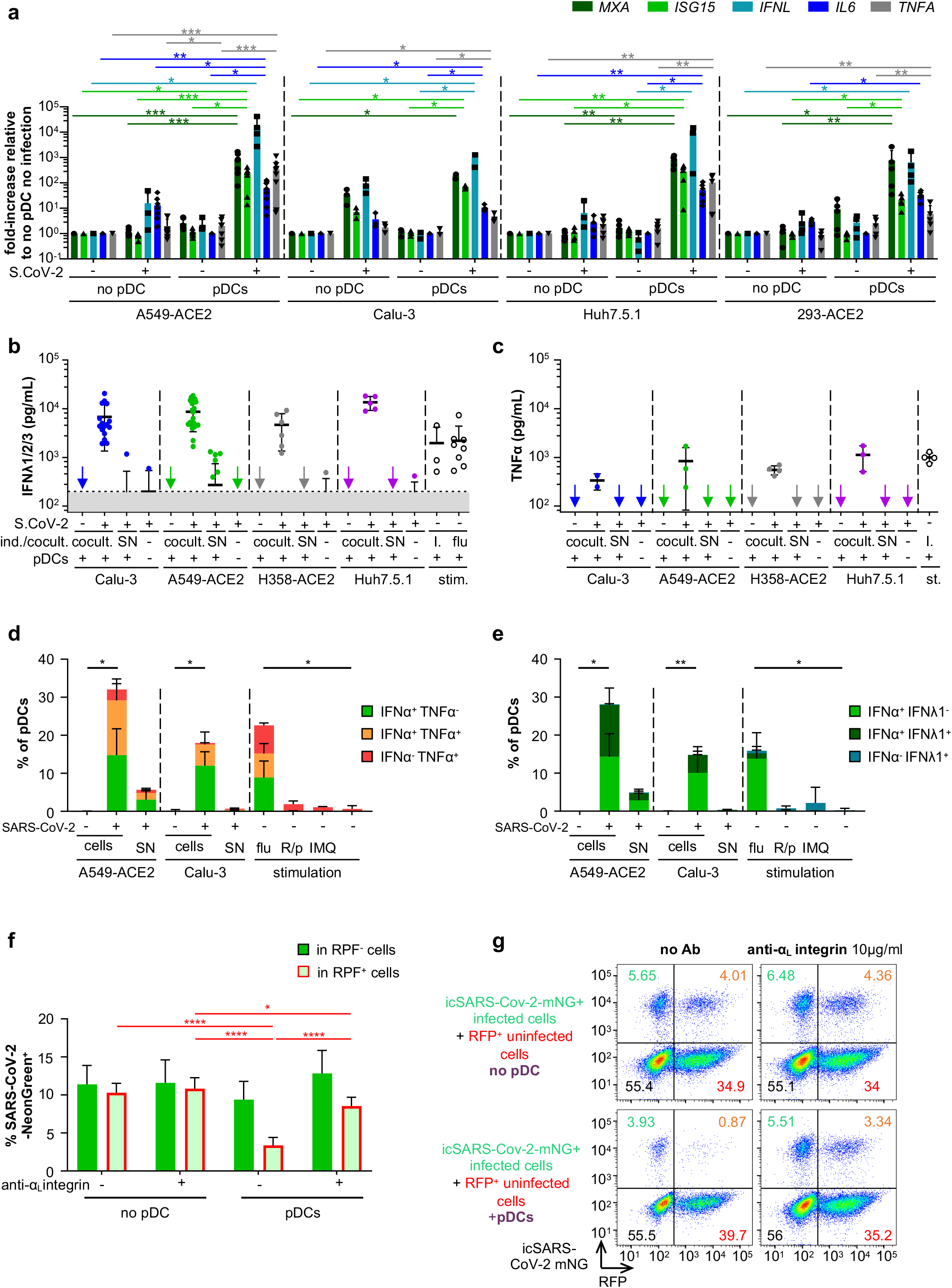
Cell-cell contact sensing of SARS-CoV-2-infected cells by pDCs induces a robust production of IFN-I/λ and other cytokines leading to the inhibition of viral spread. Human pDCs isolated from healthy donors were cocultured with SARS-CoV-2-infected (+) or uninfected (-) cells or were incubated with 100*µ*l of cell-free SN collected immediately prior to coculture from the corresponding SARS-CoV-2-infected cells, or were stimulated by influenza virus or agonists (as in **Fig. 1 and 4**). **a**, Quantification of the induction of ISG (*mxA, isg15*), type III IFN (*ifn*λ1) and pro-inflammatory cytokines (*il6, tnfA*) mRNAs in SARS-CoV-2-infected or uninfected A549-ACE2, Calu3, Huh7.5.1 or 293-ACE2 cells cultured with (pDC) or without pDC (no pDC) by RT-qPCR. Means ± SD; n=3-8 independent experiments using distinct healthy donors; statistical analysis using Kruskal-Wallis Global test; p-values as * ≤0.05, ** ≤0.005 and *** ≤0.0005 (Tukey and Kramer test). **b-c**, Quantification of IFNλ1/2/3 (**b**) and TNFα (**c**) in SN of pDCs cocultured with SARS-CoV-2-infected cells [Cocult] versus SN, as indicated. Bars represent means ± SD; n=3-14 (left panel) n=2-4 (right panel) independent experiments using distinct healthy donors. **(d-e)** Quantification by flow cytometry of the frequency of pDCs positive for IFNα and/or TNFα (**d**) and IFNα and/or IFNλ1 (**e**). Bars represent means ± SD; n=3-5 independent experiments using distinct healthy donors. **f-g**, A549-ACE2 cells were infected by icSARS-CoV-2-mNG for 24 hours and then cocultured with isolated pDCs for 48 hours. Cocultured cells were treated or not with anti-α_L_ integrin blocking antibody (10 *µ*g/mL). Viral transmission from icSARS-CoV-2-mNG+-infected cells to RFP+ uninfected cells in cocultures with pDC or without pDCs (no pDC) was quantified by flow cytometry (**f**) and representative dot plots (**g**). Results are expressed as the percentage of cells positive for mNeonGreen (mNG^+^) in the RFP^+^ (red numbers) and RFP^-^ (green numbers) cell populations. Means ± SD; n=4-5 independent experiments.

Collectively, our data provided evidence that, in contrast to stimulation with cell-free viruses and agonists, the pDC response to contact with SARS-CoV-2-infected cells is biased towards IRF7-mediated signaling that leads to a robust IFN-I/λ production.

### The pDC response controls SARS-CoV-2 spread and replication

As the response to SARS-CoV-2-infected cell primarily induced IFN-I/λ antiviral signaling, we next aimed to define how the pDC response inhibits viral propagation. pDCs were cocultured for 48 hours with icSARS-CoV-2-mNG-infected A549-ACE2 cells (mNG^+^) and uninfected A549-ACE2 RFP^+^ cells, and viral spread was quantified by flow cytometry. The results demonstrated that the pDC response readily prevented viral spread from mNG^+^ infected cells to initially uninfected RFP^+^ cells (**Fig. 4f-g**). The inhibition of contact between pDCs and infected cells *via* blockade of α_L_ integrin restored viral propagation to levels comparable to those measured in the absence of pDC (**Fig. 4f-g**). This indicated that the establishment of cell-contact *via* adhesion molecules is required for pDC-mediated antiviral response.

Interestingly, the impact of pDC antiviral response on cells infected prior to coculture (*i*.*e*., RFP^-^mNG^+^ infected cells) was lower as compared to the spread to uninfected cells (*i*.*e*., RFP^+^) (**Fig. 4f-g**). This is likely owing to the inhibition of IFN-I/λ signaling by SARS-CoV-2 within infected cells^**69,70**^. Therefore, we hypothesized that the reduction could be more potent if infected cells were directly in contact with pDCs as such contact could allow a concentrated antiviral response toward the infected cell. To test this hypothesis, we established an assay of 24 hour-long live-imaging of the coculture of pDCs (stained with CM-Dil; red) and icSARS-CoV-2-mNG infected cells (mNG^+^; green) using spinning-disk confocal microscopy. As depicted by the example of time-sequence imaging (**Fig. 5a**), pDC contact with mNG^+^ infected cells lead to a control of viral replication in the targeted infected cells, reflected by the decreased mNG fluorescent reporter signal. We controlled that these infected cells, although not mNG^+^ anymore, were still physically present by using enhanced fluorescent signal analysis (**Fig. 5a**; lower panels). Viral control by pDCs was seen for pDC/infected cell contacts starting at different time points in the course of the coculture (*i*.*e*., up to 14 hours after record onset, *data not shown*). Similar analysis performed for several infected cells, either in contact with a pDC *versus* not in contact, revealed that the decrease in viral replication (*i*.*e*., mNG fluorescence intensity) was observed only for infected cells directly in contact with pDCs (**Fig. 5b, d-e**). Of note, live-tracking of mNG fluorescence intensity performed in infected cells cultured without pDCs, provided the basal level of the mNG fluorescence intensity (done simultaneously with record of coculture with infected cells). This basal level was comparable to the one measured in infected cells cocultured with pDCs that were not in direct contact with infected cells (**Fig. 5c-d**). pDCs established long-lasting contact with infected cells *i*.*e*., mostly above 10 hours, and accordingly, the decrease in mNG fluorescence intensity was detected several hours after the onset of contact, likely owing to the time-window needed to inhibit viral replication (**Fig. 5e-f**).

**Fig. 5.**
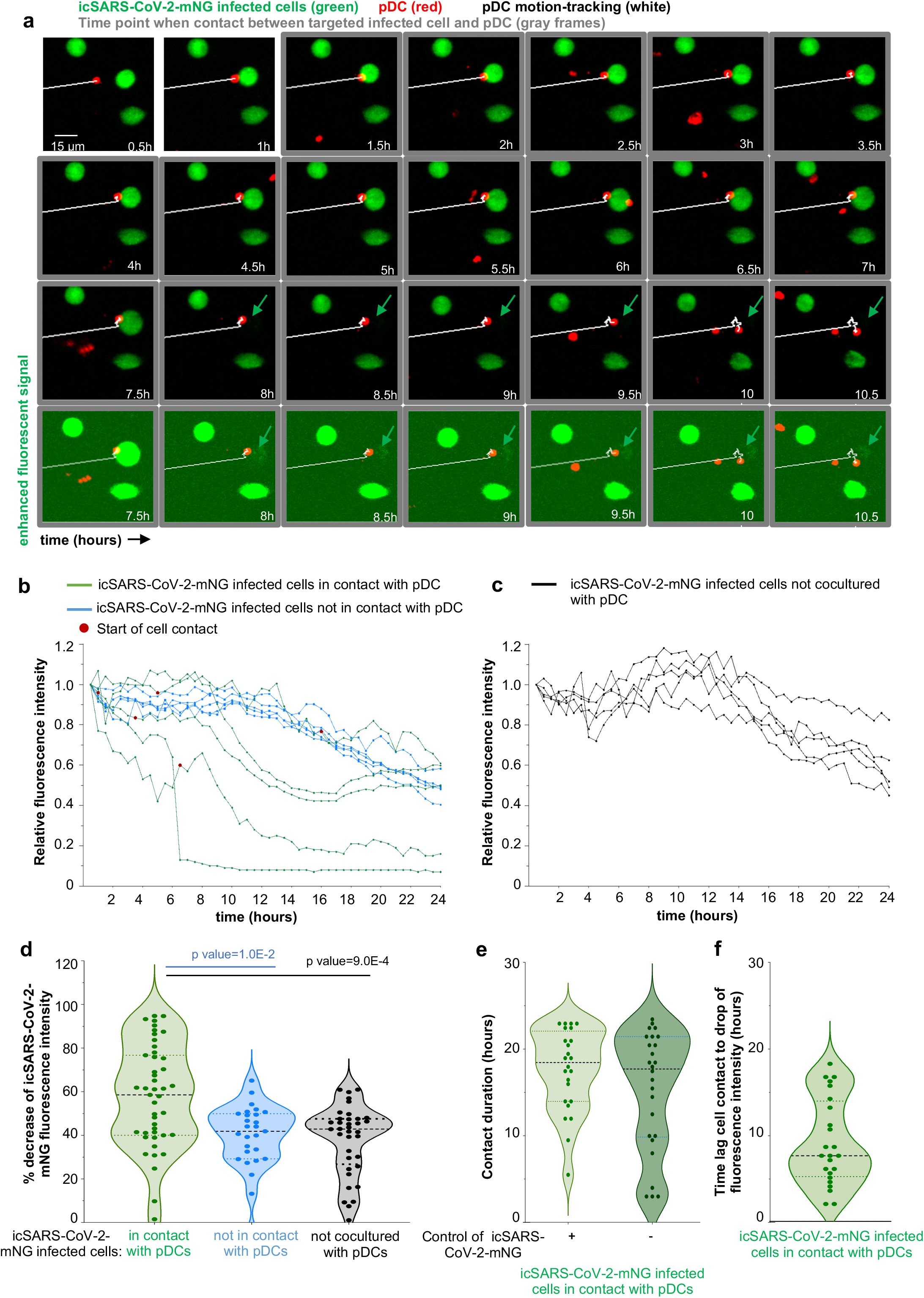
Targeted antiviral activity of pDCs toward SARS-CoV-2-infected cells. Live imaging of coculture of icSARS-CoV-2-mNG-infected cells with pDCs (CM-DiI stained) by spinning-disk confocal analysis. A549-ACE2 cells were infected by icSARS-CoV-2-mNG for 48h prior to coculture with pDCs. **a**, Representative time-sequence of pDCs (red), tracked using motion automatic tracking plug-in in image J (white line) in contact with icSARS-CoV-2-mNG-infected cells (green arrow). The time points when pDCs are in contact with infected cells are framed in grey. Bottom panels show same imaging of the seven last time points with enhanced fluorescence signal. **b-c**, Calculation of mNeongreen fluorescence intensity over time in individual icSARS-CoV-2-mNG-infected cells cocultured with pDCs: in contact (green curves) *versus* not in contact (blue curves) with pDCs (**b**), and as control/reference, in simultaneously recorded cultures of icSARS-CoV-2-mNG-infected cells without pDC (**c**), using area integrated intensity and mean value (quantification tools in Image J). The time point corresponding to the onset/start of contact is indicated by a red dot. The time point when the contact stated is indicated by red dots. The results are presented as the mNeongreen fluorescence intensity at the indicated time relative to time 0 of record set to 1; n= 5 individually recorded cells analyzed per condition from one representative experiment (and n=10-12 in other experiments). **d**, Violin plot representation of the decrease of mNeongreen fluorescence intensity (percentage) in icSARS-CoV-2-mNG-infected cells in the indicated conditions. Each dot represents one infected cell (n=118); 4 independent experiments. **e**, Violin plot representations of the contact duration between icSARS-CoV-2-mNG-infected cells and pDCs leading to control of viral replication, defined as drop of fluorescence intensity > 50% relative to the initial mNG fluoresent intensity (+) or not (-). **f**, Violin plot representations of time-lag between the onset of pDC contact and the drop of mNeongreen fluorescence intensity defined as >50% of the initial fluorescence intensity. Each dot represents one individual icSARS-CoV-2-mNG-infected cells in contact with pDCs n=50 (**e**) and n=25 (**f**) from 4 independent experiments.

Overall, these results demonstrated that pDCs established sustained contact with SARS-CoV-2 infected cells *via* α_L_β_2_ integrin/ICAM-1 adhesion complex leading to an efficient antiviral response directed toward the infected cells that shut down viral replication.

## Discussion

Here we demonstrate that pDCs are the key mediators of the IFN-I/λ response against SARS-CoV-2-infected cells. Importantly, our study of immune cells from COVID-19 patients at the single-cell and functional levels establishes that the pDC response is pivotal to control COVID-19 severity. Especially, the sensing of SARS-CoV-2-infected cells is defective in patients with severe disease. As opposed, in healthy donors the scanning function of pDCs for immune surveillance operates *via* the establishment of sustained contacts with SARS-CoV-2-infected cells *by* cell adhesion molecules. This sensing induces IRF7/IFN-I/λ-prioritized signaling in pDCs, while leaving inactive the NF-κB-mediated pathway. Next, the pDC-mediated IFN-I/λ response is specifically targeted towards SARS-CoV-2 infected cells. This specialized function thus enables pDCs to efficiently turn-off viral replication, likely owing to a concentrated efflux of antiviral effectors at the contact site with infected cells (**Fig. 6**).

**Fig. 6.**
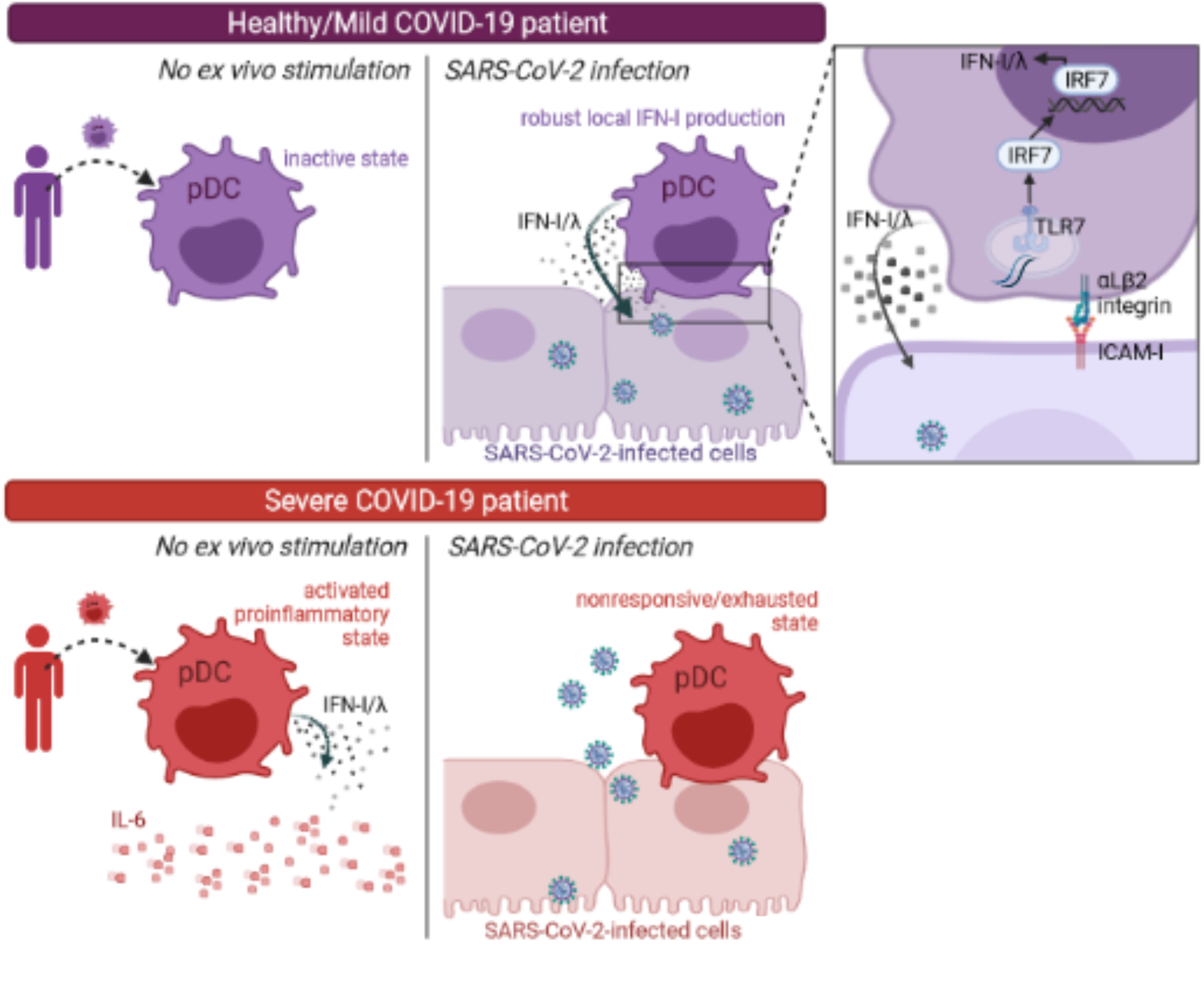
Proposed model of the innate responses associated with COVID-19 severity. Our longitudinal study of the innate responses by *ex vivo* stimulation of PBMCs from COVID-19 patients, and across distinct disease severities (*i*.*e*., *Mild*/Asymptomatic *versus Severe* COVID-19, and *healthy donors* as reference) reveals the following proposed model. (1) pDCs from *Mild* patients and *healthy donors* (in purple) robustly produce IFN-I/λ upon cell contact with SARS-CoV-2-infected cells (upper panel). (2) As opposed, pDCs from *Severe* patients (in red) produce IFN-I/λ in absence of *ex vivo* stimulation, but fail to be activated by the contact to SARS-CoV-2-infected cells (lower panel). (3) This non-responsive/exhausted state of pDCs in *Severe* COVID-19 patients is associated to an elevated level of pro-inflammatory cytokines (here represented by IL6, red round symbols) that are most likely produced by the HLA-DR^+^CD14^+^ monocytes. (4) As shown on the zoomed view of the contact site (right panel, at the top), the short-range sensing of SARS-CoV-2-infected cells by pDCs requires cell contact mediated by adhesion complexes, identified as α_L_β_2_ integrin and ICAM-1. This triggers a TLR7-induced signaling *via* IRF7 leading to an IFN-I/λ-prioritized response, while leaving inactive the NF-κB-mediated signaling.

### pDCs critically control viral replication and the outcome of COVID-19 severity

Insights on several hallmarks of IFN-I/λ pathway as being critical in COVID-19 severity are emanating from recent publications. First, neutralizing autoantibodies against several cytokines and genetic defects affecting the IFN-I pathway have been identified in life-threatening COVID-19, as autosomal disorders of IFN-I immunity and autoantibodies underlie at least 10% of critical COVID-19 pneumonia cases^**21-27**^. Second, patients with severe diseases exhibit reduced circulating pDCs and low plasma IFN-I/λ levels when compared to mild COVID-19 patients^**7,19,20**^. Third, IL-3, which increases innate immunity likely by promoting the recruitment of circulating pDCs into the airways, is reduced in the plasma of patients with high viral load and severity/mortality^**71**^. This evidence highlights that several biomarkers of IFN-I/λ response are diminished in COVID-19 patients, and hereby points to pDCs as a primary candidate in the progression of COVID-19. Here, using an original approach to study innate immunity in *ex vivo*-stimulated PBMCs from COVID19 patients, we further report that the functionality of pDCs is markedly blunted in severe patients, as observed upon their stimulation by SARS-CoV-2-infected cells as well as other TLR agonists.

### pDC effector functions against SARS-CoV-2

pDCs comprise distinct subpopulations capable of varied functions and efficacy levels to mount an IFN-I/λ response^**28,34,60-63-72-73**^. Viral infections and attendant inflammation potentially impact the frequency and functionality of the distinct pDC subpopulation including effect on pDC renewal^**60,72,74**^. Such modulations could be imprinted either by the micro-environment *via* crosstalk with immune cells, or by pDC activation itself.

We now report that neither pDC subpopulation frequencies nor their distinct ability to respond to SARS-CoV-2 are impacted by stimulation and contact with SARS-CoV-2-infected cells. This is notably illustrated for CD2^low^/CD2^hi^ pDCs that display similar activation profile upon stimulation with SARS-CoV-2-infected cells.

Interestingly, when comparing stimulations by contact with SARS-CoV-2-infected cells *versus* cell-free activators (*i*.*e*., viral particles and synthetic agonists), we found that the differentiation of pDCs defined by CD80 and/or PD-L1 expression are readily distinguishable depending on the TLR7 inducers. Whilst pDCs diversify towards all the different PD-L1/CD80 subsets upon cell-free stimulation (agonist and virus) in agreement with prior publications^**34,60**^, the contact with infected cells restricts their evolution to PD-L1^+^CD80^-^ subsets. Of note, previous reports suggested that PD-L1^+^CD80^-^ pDCs are more efficient for IFN-I response compared to the other subsets^**34,60,72**^.

In accordance with this phenotype, the contact with SARS-CoV-2-infected cells induces a IRF7/IFN-I/λ-prioritized signaling in pDCs, while leaving inactive the NF-κB-mediated pathway. The ‘bifurcated’ signaling in TLR7-activated pDCs can occur independently toward either IFN-I/λ production or NF-κB activation leading to pro-inflammatory cytokines and activation markers. This bifurcation of signaling is expected to be dependent on the sub-cellular compartment in which the TLRs encounter activating signal: activation in early endosomes induces IFN-I while activation in endolysosomes triggers NF-κB-mediated signaling^**28,75**^. We previously reported that, in the context of other viral infections (*i*.*e*., Dengue, Chikungunya, Hepatitis C, Zika viruses), contact with infected cells similarly induced a IRF7/IFN-I/λ prioritized signaling in activated pDCs, as opposed to incubation of pDCs with cell-free viruses^**47,68**^. Of note, we also show that stimulation of pDCs by cell-free SARS-CoV-2 supernatants induced the upregulation of some activation markers (*e*.*g*., HLA-DR, PD-L1), in accordance with other reports^**34,60**^, but elicited virtually no IFN-I/λ response. We propose that the signaling downstream of TLR7, including the phosphorylation cascade might be impacted by cell polarity and physical contact with infected cells^**76,77**^. Further studies will aim at addressing this question.

### Cross-regulation of immune responses

Progression to severe COVID-19 predominates in elderly patients, since advanced age is a factor suspected to aggravate disease progression and to weaken innate immunity, potentially including pDC responsiveness^**78,79**^. This is in accordance with the demographic analysis of our COVID-19 cohort, as the age-ranges were 63.5-year-old [interquartile of 48.0-73.0] and 40.5-year-old [24-57] for *Severe* and *Mild* patients, respectively. The patient history and genetic factors can also explain the differential ability of pDCs from patients to mount a response against SARS-CoV-2-infected cells, as illustrated by the heterogeneous immune responses of patients, with critical anomalies in functionality of pDC in severe COVID-19. Our kinetic analysis performed on a COVID-19 patient that presents auto-antibodies against IFN-I^**21-23-25**^ shows that the responsiveness of their pDCs to stimulation by SARS-CoV-2-infected cells was blunted **(Fig. 2i)**. A future study is needed to expand this interesting preliminary observation, which already indicates that the IFN-I/λ response can reinforce via a positive feedforward regulation the pDC antiviral function.

Of note, comparison of responses across patients with different COVID-19 severities suggest that pDC response to SARS-CoV-2-infected cells inversely correlates with an exacerbated inflammatory response (as illustrated by IL6 production by monocytic cells) and a basal level of IFN-I/λ and inflammation activity. It is tempting to speculate that the progressive enrichment of proinflammatory cytokines in the lung micro-environment can imprint pDC responsiveness. In this scenario, an excessive elicitation of pDCs would lead to their functional ‘silencing’. Future studies will be needed to elucidate the underlining mechanism that could lead to such ‘*exhausted*’ pDCs, as reminiscent of the findings of impaired pDC function in a distinct infection context^**74**^. In turn, the deficit of the antiviral control at the infected site owing to *exhausted* pDCs can then feedback as fuel for uncontrolled viral replication leading to more lung inflammation. Altogether our results thus highlight possible cross-regulation between immune cells in the course of COVID-19.

### Concluding remarks: importance for SARS-COV2 and possible insight for predictive markers

SARS-CoV-2 is a still-ongoing worldwide health threat, currently causing a significant human and economic burden, being exacerbated by divergence into more severe variants. Here, we provide compelling evidence that pDCs are a key cell type in the initiation of antiviral responses against SARS-CoV-2. Further, this study identified the failure of pDC response as critical in COVID-19 severity. Moving forward our finding shall provide guidelines for predictive biomarkers, as associated with pDC responsiveness to SARS-CoV-2 infections and background IFN-I/λ response at different stages of disease. Furthermore, strategies to boost the pDC response, and especially their recruitment to the lung, can lead to the development of potential therapeutics against pulmonary viral infections.

## Methods

### Preparation of Viral Stocks and Infections

The clinical isolate was obtained from patients referenced in the GISAID EpiCoVTM database: as BetaCoV/France/IDF0571/2020; accession ID: EPI_ISL_411218^**80**^, kindly provided by Dr B. Lina. The infectious-clone-derived mNeonGreen SARS-CoV-2 (referred to as icSARS-CoV-2-mNG) was kindly provided by Dr Pei-Yong Shi and generated by introducing mNeonGreen into ORF7 of the viral genome^**45**^. Viral stock of Influenza A Virus (Flu A/H1N1/New caledonia ; infectious titer of ≈ 10^7^ plaque forming unit (PFU)/ml)^**81**^ was produced as previously described and kindly provided by Dr V. Lotteau (CIRI, Lyon France).

### Cell Lines and Primary Cell Cultures

SARS-CoV-2-infected cells included the human alveolar basal epithelial cell lines, Calu-3 cells (ATCC HTB-55), A549 cells (ATCC CCL-185) and NCI-H358 cells (ATCC CRL-5807), Huh7.5.1 cells^**82**^ and HEK-293 cells (ATCC CRL-1573). The A549 cells, NCI-H358 cells and HEK-293 cells were transduced to stably express the human angiotensin converting enzyme 2 (ACE2; accession number: NM_021804) using a lentiviral vector, as previously described^**82,83**^. A549 cells were maintained in Roswell Park Memorial Institute (RPMI) 1640 Medium (Life Technologies) supplemented with 10% FBS, 100 units (U)/ml penicillin, 100mg/ml streptomycin and non-essential amino acids (Life Technologies) at 37°C/5% CO_2_. The NCI-H358 and Huh7.5.1 cells were maintained in Dulbecco’s modified Eagle medium (DMEM) (Life Technologies) with the same supplements and 2mM L-glutamine. Calu-3 cells were maintained in DMEM/Nutrient mixture F-12 Ham (1:1) (Life Technologies) supplemented with glutaMAX, 10% FBS, 100 units (U)/ml penicillin, and 100mg/ml streptomycin. All cell were maintained at 37°C/5% CO_2_.

pDCs were isolated from 450 ml of blood units obtained from adult human healthy donors and according to procedures approved by the “Etablissement Français du sang” (EFS) Committee. PBMCs were isolated using Ficoll-Hypaque density centrifugation. pDCs were positively selected from PBMCs using BDCA-4-magnetic beads (MACS Miltenyi Biotec) and cultured as previously described^**47**^. PBMCs and pDCs were cultured in RPMI 1640 Medium (Life Technologies) supplemented with 10% FBS, 100 units (U)/ml penicillin, 100 mg/ml streptomycin, 2 mM L-glutamine, non-essential amino acids, 1 mM sodium pyruvate and 10 mM Hepes (Life Technologies) at 37°C/5% CO_2_.

### Cohort of SARS-CoV-2 infected patients

The constitution of the cohort was done by the collaboration of the Hospices Civils de Lyon (HCL). This cohort consists of patient groups recognized by clinicians as: *i)* patients admitted in intensive care units for severe disease at hospital admission (*i*.*e*., acute respiratory distress syndrome or severe pneumonia requiring mechanical ventilation, sepsis and septic shock) are referred to as *Severe group* and *ii)* patients with mild symptoms (*i*.*e*., low-grade fever, cough, malaise, rhinorrhea, sore throat) are referred to as *Mild early group* when collected in the first two weeks and *Mild late group* for later time points. The detailed description of the patient information also includes the levels of the IFN-I/III signature and viremia, as determined in blood and nasal swab samples, respectively (*data not shown*). As reference, blood samples from *healthy donors* were obtained from EFS and experimentally processed similarly. To limit the risk of inclusion of asymptotic healthy donor: *i/* part of the blood samples was collected prior to the pandemic and *ii/* for blood collected during the SARS-CoV-2 pandemic, systemic examination and questioning/interview of the donors were performed and included symptoms, prior contacts at risk and vaccination, Thus, blood samples were excluded from our study if ongoing and/or recent COVID-positivity was suspected.

### Reagents

Ficoll-Hypaque (GE Healthcare Life Sciences). Other reagents included LPS, TLR3 agonist (Poly(I:C); LMW) and TLR7 agonist (R848 and Imiquimod) (Invivogen); TLR7 antagonist (IRS661, 5’-TGCTT GCAAGCTTGCAAGCA-3’ synthesized on a phosphorothionate backbone; MWG Biotech); mouse anti-α_L_ integrin (clone 38; Antibodies Online); mouse anti-ICAM-1 (Clone LB-2 ; BD Bioscience): Arp2/3 complex inhibitor I (CK-666 ; Merck Millipore) ; Fc Blocking solution (MACS Miltenyi Biotec); Golgi-Plug, cytoperm/cytofix and permeabilization-wash solutions (BD Bioscience); IFNα and IFN*λ1/2/3* ELISA kit (PBL Interferon Source); IL6 and TNFα ELISA kit (Affymetrix, eBioscience); 96-well format transwell chambers (Corning); IL6 and IFN*λ* by U-PLEX Custom Human Cytokine assay (Meso Scale Diagnostics, Rockville, MD) ; 96-Well Optical-Bottom Plates (Thermo Fisher Scientific); cell-labeling solution using CellTraceTM Violet Cell Proliferation Kit (Life Technologies ref # C34557, C34571), Live/Dead Fixable Dead Cell Stain Near-IR (Life Technologies ref #10119); Fixable Viability Dye eFluor 450 (Life Technologies); Zombie Aqua and Zombie Green Fixable Viability Kits (Biolegend); FITC Annexin V Apoptosis Detection Kit with 7-AAD (Biolegend); cDNA synthesis and qPCR kit (Life Technologies) ; poly-L-lysin (P6282, Sigma-Aldrich).

### Quantification of SARS-CoV-2 level in nasal swab samples of infected patients

Nucleic acid extraction was performed from 0.2 mL naso-pharyngeal swabs using NUCLISENS easyMAG and amplification was performed using Biorad CFX96. Quantitative viral load was determined using four internally developed quantification standards (QS) targeting the SARS-CoV-2 N gene: QS1 to QS4 respectively at 2.5.10^6^, 2.5.10^5^, 2.5.10^4^, 2.5.10^3^ copies/mL of a SARS-CoV-2 DNA standard. These QS were controlled and quantified using the Nanodrop spectrophotometer (ThermoFisher) and Applied Biosystems QuantStudio 3D Digital PCR. In parallel, naso-pharyngeal swabs were tested using the CELL Control R-GENE kit (amplification of the HPRT1 housekeeping gene) that contains two quantification standards QS1 and QS2, at 10^4^ copies/*µ*L (50,000 cells/PCR *i*.*e*. 1.25.10^6^ cells/mL in our conditions) and 10^3^ copies/*µ*L (5000 cells/PCR *i*.*e*. 1.25.10^5^ cells/mL in our conditions) of DNA standard, respectively, to normalize the viral load according to the sampling quality.

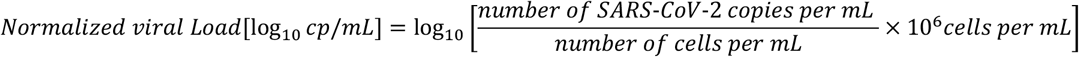

### Analysis of IFN-I/λ signatures in blood samples of SARS-CoV-2-infected patients

#### Transcript levels using Nanostring technology

Targeted transcripts of the IFN-I pathway included SIGLEC1, IFI27, IFI44L, IFIT1, ISG15, RSAD2, HPRT1, POLR2A and ACTB^**84**^. Transcript levels were determined using RNA extracted from the patient’s blood samples by Maxwell 16 LEV simply RNA Blood kits that comprised an individual DNAse treatment^**84**^. Next, total RNA was eluted in 40 *µ*L RNAse-free water and concentration was quantified by spectrophotometry using a NanoVue (Biochrom). Only 200 ng of RNA were needed to achieve IFN signature with Nanostring technology. For the Elements system, capture (probe A) and tag (probe B) probe DNA oligos were designed by NanoString and synthesized by Integrated DNA Technologies (IDT). After hybridization at 67°C for 18-20 hours, samples were analyzed using the “High Sensitivity” protocol option on the nCounter Prep Station and counted on nCounter Digital Analyzer using maximal data resolution. Data were processed with nSolver software (NanoString Technologies, Seattle, WA), which included assessment of the quality of the runs, and combined, normalized, and analyzed in nSolver and Excel.

Normalization was performed by applying a scaling factor that normalizes the geometric mean of housekeeping genes (β-Actin; *ACTB*, hypoxanthine phosphoribosyltransferase 1; HPRT1 and RNA polymerase II subunit A ; POLR2A) for each sample and these normalized counts were used to calculate the scores, as previously reported^**84**^. The median of these ISG relative expression was used as an IFN score.

#### IFNα protein measurement by Simoa technology

All reagents were purchased from Quanterix (reference 100860) and loaded onto the Simoa HD-1 Analyzer (Quanterix) according to the manufacturer’s instructions and using three-step assay configurations. Briefly, the beads were pelleted with a magnet to remove supernatant (SN). Following several washes, 100 uL of detector antibody were added, according to the manufacturer’s instructions. The beads were then pelleted with a magnet, followed by washes and 100μL of ß–D–galactosidase (SβG) were added. The beads were washed, re-suspended in resorufin ß-D-galactopy-ranoside (RGP) solution, and loaded onto the array. The array was then sealed with oil and imaged. Images of the arrays were analyzed and AEB (average enzyme per bead) values were calculated by the software in the HD-1 Analyzer, as previously reported^**85**^. Human plasma samples along with calibration curves were measured using the Simoa HD-1 Analyzer. The calibration curves were fit using a 4PLfit with 1/y2 weighting factor and were used to determine the concentrations of the unknown human plasma samples. This analysis was done automatically using the software provided by Quanterix with the Simoa HD-1 Analyzer.

#### IL6, IFNλ1 and IFNγ protein measurements

Concentrations were determined in patient’s serum using U-PLEX Custom Human Cytokine assay (Meso Scale Diagnostics, Rockville, MD). The assays were performed according to the manufacturer’s instructions with overnight incubation of the diluted samples and standards at 4°C. The electrochemiluminescence signals (ECL) were detected by MESO QuickPlex SQ 120 plate reader (MSD) and analyzed with Discovery Workbench Software (v4.0, MSD).

### *Ex-vivo* stimulation of PBMCs isolated from SARS-CoV-2-infected patients and healthy donors

Bloods of SARS-CoV-2-infected patients and healthy donors were collected in EDTA tubes. PBMCs were freshly isolated by Ficoll-Hypaque density centrifugation followed by washing in pDC/PMBC culture medium (*i*.*e*., RPMI 1640 Medium supplemented with 10% FBS, 100 U/ml penicillin, 100 mg/ml streptomycin, 2 mM L-glutamine, non-essential amino acids, 1 mM sodium pyruvate and 10 mM Hepes). PMBCs were frozen in 1mL freezing medium (10% DMSO, 90% FBS) and cryopreserved in vapor phase liquid nitrogen (>-135°C). Two hours prior to *ex vivo* stimulation, PBMCs were thawed out at 37°C rinsed with 10 mL of FCS, incubated in 40 mL of pDC/PMBC culture medium at 37C°/5% CO_2_ for 30 minutes and resuspended in culture medium. 2.5×10e5 PBMCs were cocultured with 1×10e5 SARS-CoV-2-infected *versus* uninfected A549-ACE2 cells, as negative control, or were stimulated with TLR agonists [31.8 *µ*M R848 and 42.22 *µ*M polyI:C] for the FACS panel of mDC1/pDC analysis and LPS stimulation [2.04 *µ*M] for the mDC2/non-mDC2/HLA-DR^+^ CD14^+^ panel in a final volume of 200*µ*l in 96-well round-bottom plates incubated at 37C°/5% CO_2_ for 14 to 16 hours. Cell-culture SNs were collected for quantification of cytokine levels (IFNα by ELISA/Simoa; IL6, IFN and IFNλ1 by U-PLEX Custom Human Cytokine assay) while cells were harvested for flow cytometry or for Nanostring analyses.

RNAs were isolated from these cells by phenol/chloroform extraction procedure as previously described^**47**^. The subsequent steps of the procedure used for Nanostring analysis on *ex-vivo* stimulated PBMCs were performed as described above for the patient blood samples.

### Coculture experiments using isolated pDCs

Unless indicated differently, 2×10e4 pDCs were cocultured with SARS-CoV-2-infected or uninfected cells as 5×10e4 or 1×10e5 cells for analysis by RT-qPCR or flow cytometry, respectively or were stimulated with 100*µ*l of cell-free SN collected from SARS-CoV-2-infected cells. The cells were infected at MOI 0.01, 0.1, 0.02 and 0.5 for, respectively, NCI-H358-ACE2, Huh7.5.1, A549-ACE2 and Calu-3 cells for 48 hours maximum prior to collection of the cells and their SNs for coculture. As comparison pDCs were stimulated with TLR agonists [31.8 *µ*M R848 and 42.22 *µ*M polyI:C] in a final volume of 200*µ*l in 96-well round-bottom plates incubated at 37C°/5% CO_2_. When indicated, cells were cocultured in 96-well format transwell chambers (Corning) with a 0.4 *µ*m permeable membrane. At the indicated time, cell-culture supernatants were collected for quantification of cytokine levels: IFNα, IFN*λ1/2/3* (IL29/28A/28B), TNFα and IL6 using specific ELISA kit (PBL Interferon Source, Affymetrix, respectively) following the manufacturer’s instructions. Cells were harvested at the indicated times for analysis by flow cytometry or RT-qPCR.

### Immunostaining and flow cytometry analysis

At the indicated times, harvested cells were resuspended using 2 mM EDTA-PBS solution for the coculture with PBMCs and 0.48 mM EDTA-PBS solution for pDC cocultures. Cells were incubated with 1 μL/mL viability marker diluted in PBS for 20 minutes at RT. After a 10-minute incubation with Fc receptor blocking reagent (MACS Miltenyi Biotec) at 4°C followed by two PBS washes, cells were stained for surface markers for 30 minutes at 4°C with antibodies diluted in staining buffer (PBS without calcium and magnesium, with 2% FBS and 2mM EDTA), followed by two PBS washes. These markers included generic lineage markers (CD3, CD19, CD20, CD56 for exclusion, and CD11c, HLA-DR for selection of cell populations), and specific markers of pDCs (CD123, BDCA-2, CD2 and Axl), mDC2 (BDCA-1), mDC1 (BDCA-3), monocytes (CD14 and CD16) and/or cell differentiation markers (CD83, CD80 and PD-L1). For the identification of apoptotic and necrotic cells, surface-stained cells were labelled using FITC Annexin V Apoptosis Detection Kit with 7-AAD according to the manufacturer’s instructions. Following one wash with Annexin V Binding Buffer (Biolegend), cells were fixed with 4% PFA for 30 minutes at 4°C. For intracellular-immunostaining, cells were treated with 1 *µ*l/ml GolgiPlug solution (BD Bioscience) for 3 hours at 37°C/5% CO_2_ before collection. After surface staining and fixation with cytoperm/cytofix solution (BD Bioscience) for 20 minutes at 4°C, IFNα, IL6, IFN*λ1*, and TNFα were stained by a 45-minute incubation at 4°C with antibodies diluted in permeabilization buffer (BD Bioscience). Cells were then washed with permeabilization buffer and resuspended in staining buffer. Flow cytometric analysis was performed using a BD LSR Fortessa 4L. Compensation beads were used as reference for the analysis. The data were analyzed using Flow Jo software (Tree Star).

### Analysis of transcriptional levels by RT-qPCR

RNAs were isolated from cells harvested in guanidinium thiocyanate citrate buffer (GTC) by phenol/chloroform extraction procedure as described previously^**47**^. The mRNA levels of human *MXA, ISG15, IFNL, IL6, TNFA* and glyceraldehyde-3-phosphate dehydrogenase (*GADPH*) were determined by RT-qPCR using iScript RT kit (Life Technologies) and PCR Master Mix kit (Life Technologies) for qPCR and analyzed using StepOnePlus Real-Time PCR system (Life Technologies). The mRNA levels were normalized to *GADPH* mRNA levels.

### Analysis of extracellular infectivity

Infectivity titers in supernatants were determined by end-point dilution in plaque assay. Briefly, 10-fold serial dilutions of SARS-CoV-2-containing supernatants were added to 2×10e5 Vero cells seeded in 12-well plates for a 2 hour-incubation. The medium was then replaced by DMEM containing 2% FBS and 2% carboxymethylcellulose (CMC). The cytopathic effect was scored 96 hours post-infection : cells were fixed for 30 minutes with 4% PFA and colored by cristal violet solution.

For icSARS-CoV-2-mNG infection, foci were directly detected according to mNeongreen-positive cells. Briefly, 10-fold serial dilutions of icSARS-CoV-2-mNG-containing supernatants were added to 2×10e4 Vero cells seeded in 96-well plates and fixed 24 hours post-infection. GFP-expressing cells were quantified by foci counting using a Zeiss Axiovert 135 microscope.

### Viral Spread Assay

A549-ACE2 cells were transduced with lentiviral-based vector pseudotyped with VSV glycoprotein to stably express RFP, as previously reported^**47**^. After immuno-isolation, pDCs were stained with CellTrace Violet Cell Proliferation kit (Life Technologies) for 20 minutes at 37°C in the dark. Labeled pDCs were then spinned down and resuspended in pDC culture medium. 2.5×10e4 pDCs were cocultured with 2.5×10e4 icSARS-CoV-2-mNG-infected cells (infected for 24 hours prior to coculture) and with 2.5×10e4 RFP^+^ uninfected cells for 48 hours at 37°C/5% CO_2_. When indicated, the cocultures were treated with an anti-α_L_ integrin blocking antibody at 10mg/mL. After coculture, harvested cells were stained with Live/Dead Fixable Dead Cell Stain Near-IR marker for 30 minutes at RT, washed with PBS and fixed with 4% PFA for 30 minutes at 4°C. The level of viral spread from icSARS-CoV-2-mNG-infected cells (mNG^+^) to uninfected cells (RFP^+^) during coculture was determined by flow cytometric analysis as the frequency of infected cells (mNG^+^/RFP^+^ population) among the RFP^+^ cell population and similarly in RFP^-^ populations. Flow cytometric analysis was performed using a BD LSR Fortessa 4L and the data were analyzed using Flow Jo software (Tree Star).

### Live imaging of coculture with spinning-disc confocal microscopy analysis

A549-ACE2 cells were infected with icSARS-CoV-2-mNG for 48 hours prior to coculture with pDCs. Infected cells were seeded (2×10e4cells per well) in a 96-Well Optical-Bottom Plate pre-coated with poly-L-lysine (1 hour incubation at 37°C/5% CO_2_ with 8mg/mL poly-L-lysine). Isolated pDCs were stained with 0.5*µ*M Vybrant cell-labeling solution (CM-DiI, Life Technologies) by successive incubations for 10 and 15 minutes at 37°C and 4°C respectively. After addition of pDCs to seeded infected cells, the cocultures were imaged every 30 minutes during 24 hours at magnification x10 with a BSL3-based spinning-disc confocal microscope (AxioObserver Z1, Zeiss). The cocultures were maintained at 37°C/5% CO_2_ in an incubation chamber. Analyses of pDC motion were performed using projection of Z-stacks with maximal intensity (*i*.*e*., about 5-to-10 selected Z-stacks per fields out of 30 Z-stacks in total). The quantification of mNeonGreen fluorescence intensity of infected cells and the duration of contacts between pDCs and infected cells were performed using Image J software package (http://rsb.info.nih.gov/ij). The calculations of pDC positions were performed using Trackmate plug-in of Image J software.

### PCA Analysis

PCA analyses were performed using the R princomp function (R version 4.1.0) with the correlation matrix approach^**86-89**^. All time points available for each patient with a complete dataset for any given cell type were used. The time points of the patients presenting a mild severity were discriminated between *Mild early* (≤ 15 days after estimated primo-infection) and *Mild late* (> 15 days). For each cell type, PCA analyses were performed independently for each stimulation condition with the parameters presented in the vectors plots.

### Bioinformatic analysis of the cis-acting regulatory element in the promoter

For each candidate gene, we recovered the genomic nucleotide sequence spanning from 1500 nucleotides upstream, until 100∼200 nucleotides downstream of the annotated Refseq transcription start site. For genes with more than one annotated transcription start site, we collected the sequence from 1500 nucleotides upstream of the most 5’ transcription start site until 100∼200 nucleotides downstream of the most 3’ transcription start site. The bioinformatic analyses were performed using the FIMO and AME tools available on https://meme-suite.org/meme/^**90,91**^ and the consensus sequence as follows:

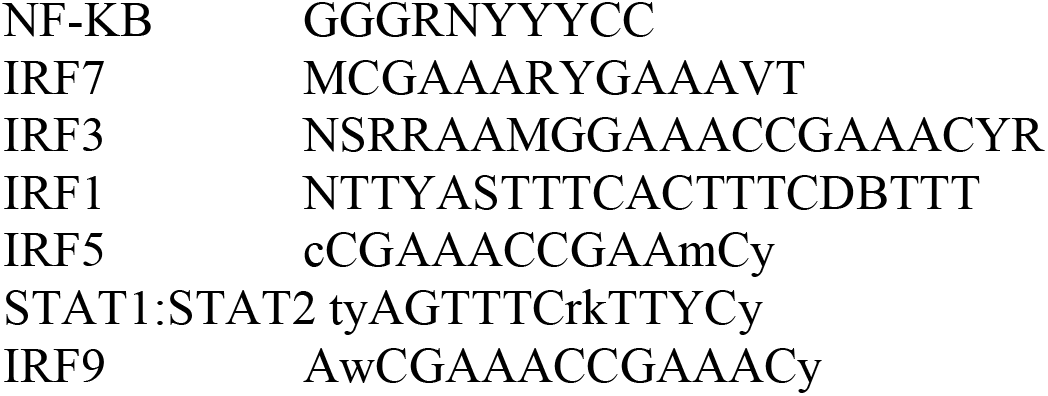

### Statistical methods

Statistical analysis was performed using R software environment for statistical computing and graphics, version 3.3.2.

For quantifications by ELISA, RT-qPCR and flux cytometry analyses of the levels of cytokines, ISG and cell surface markers, the statistical analyses were performed using one-way ANOVA on ranks (Kruskal−Wallis rank sum test). When the test was considered significant (p-values ≤ 0.05), we used the Tukey Kramer (*Nemenyi*) pairwise test as *post hoc* test for multiple comparisons of mean rank sums to determine which contrasts between individual experimental condition pairs were significant.

Of note, for each independent experiment preformed using PBMCs isolated from SARS-CoV-2 infected patients, the same procedures and analyses were done in parallel using different *healthy* donors, as references. Likewise, all independent experiments of cocultures with PBMCs or pDCs were performed using distinct *healthy* donors as reference. To test difference between the patient groups in the Flow cytometry analysis of the biomarker the different cell population, we used the Beta regression model with the logit link function from the R ‘betareg” package, which is the suitable statistical approach for modeling continuous response Y variables that vary in the open standard unit interval (0, 1). Beta regression being unable to model in case Y contains exactly 0 or 1 values, Y data was transformed to Y’ as following : Y’ = (Y * (n − 1) + 0.5)/n where n is the number of patients in all compared groups.

For the quantification by flow cytometry analysis of the viral spread of the icSARS-CoV-2-mNG SARS-CoV-2 molecular clones from RFP^-^ cells to RFP^+^ cells (initially uninfected) (**Fig. 4**) and for quantification of data from the live-imaging analysis using spinning-disk confocal microscopy analysis (**Fig. 5**), the statistical analyses were performed using one-way ANOVA followed by Tukey multiple comparisons of means.

The set of Figures was prepared using PRISM software.

## Data Availability

all data referred to in the manuscript will be available

## Author contributions

MV, MSR, ED, AliB, GJ, HP, AleB, SA and MD for conception or design of the work; MV, MSR, ED, AliB, GJ, MV, DC, MP, RP, HP, TW, OA, AleB, SA, ER and MD for acquisition, analysis, or interpretation of data; DC and ER for creation new tools used in the work; MV, MSR, ED, AliB, GJ, ER and MD have drafted the initial manucript, MV, MSR, ED, AliB, GJ, TW, AleB, ER and MD for edition of the mansucript

## Acknowledgments

We thank Dr B. Lina (CIRI, Lyon, France) for kindly provided the clinical isolate of SARS-CoV-2 ; Dr Pei-Yong Shi (University of Texas Medical Branch, Galveston, US) for the infectious clone of SARS-CoV-2 with mNeonGreen reporter; Dr V. Lotteau (CIRI, Lyon, France) for viral stock of the influenza A virus; Dr C. Goujon (Institut de Recherche en Infectiologie de Montpellier, IRIM, France) for the ACE2-lentiviral construct and Dr F.V. Chisari (Scripps Research Institute, La Jolla, CA) for the Huh7.5.1 cells. We are grateful to Drs Y. Jaillais, A Marçais, B. Webster, S Assil, P.Y Lozach and A. Bosseboeuf for critical readings of the manuscript and to our colleagues for their encouragement and help. We acknowledge the contribution of SFR Biosciences (UMS3444/CNRS, US8/Inserm, ENS de Lyon, UCBL) including the PLATIM and AniRA-cytometry facilities, especially J. Brocard and S. Dussurgey, for technical assistance for imaging and FACS analyses, respectively. We acknowledge the contribution of the EFS Confluence/Decine-Lyon. This work was supported by grants from the *Agence Nationale de la Recherche* (ANRJCJC-iSYN); the *Fondation pour le recherche médicale* (FRM; ANR Flash COVID-19); the *Agence Nationale pour la Recherche contre le SIDA et les Hépatites Virales* (ANRS – N21006CR and N19017CR); the *UDL/ANR IA ELAN ERC* (G19005CC); and *FINOVI* (AO11 – collaborating project); from EU H2020 ZIKAlliance. PhD fellowships for M.R. and G.J. are respectively sponsored by ANRS and ‘*Contrats doctoraux Lyon 1 dédiés à l’International’* from Université Lyon 1.

